# Adiposity and inflammation mediate altered metabolic profiles in individuals with opioid use disorder

**DOI:** 10.64898/2026.04.13.26350800

**Authors:** Xinyi Li, Peter Manza, Gene-Jack Wang, Natasha Giddens, Annabelle Belcher, Melanie Schwandt, Nancy Diazgranados, Kevin G. Lynch, Nora D. Volkow, Zhenhao Shi, Corinde E. Wiers

**Affiliations:** Center for Studies of Addiction, Department of Psychiatry, Perelman School of Medicine, University of Pennsylvania, Philadelphia PA; National Institute of Alcohol Abuse and Alcoholism, National Institutes of Health, Bethesda MD; Division of Child and Adolescent Psychiatry, Donald and Barbara Zucker School of Medicine, Hofstra University, Glen Oaks, NY, USA; Department of Psychiatry, University of Maryland, Baltimore MD

**Keywords:** metabolism, inflammation, opioid use disorder, nutrition, body weight gain

## Abstract

Previous studies have linked opioid use to altered metabolic profiles, but findings have been inconsistent and mechanisms remain unclear. One potential mechanism involves increased adiposity, leading to chronic low-grade inflammation that elevates metabolic risk. Here, we examined metabolic profiles in individuals with opioid use disorder (OUD) and matched non-OUD controls, focusing on the sequential mediating roles of BMI and inflammation. Data from individuals with OUD (n=281) and non-OUD (n=246) were drawn from a natural history screening protocol from the National Institute on Alcohol Abuse and Alcoholism intramural program. Groups were matched on age, sex, race, ethnicity, socioeconomic status, and education via propensity score matching. Metabolic measures included BMI, blood glucose, hemoglobin A1c (HbA1c), and lipid profiles, with lipid imbalance indexed by the atherogenic index of plasma (AIP). Inflammatory markers included C-reactive protein (CRP) and erythrocyte sedimentation rate (ESR). Individuals with OUD had significantly higher BMI (F_1,481_=12.9, p<0.001), HbA1c (F_1,481_=10.5, p=0.001), lower high-density lipoprotein cholesterol (HDL-C; F_1,481_= 46.2, p< 0.001), higher low-density lipoprotein cholesterol (LDL-C; F_1, 481_=11.9, p< 0.001), and higher AIP (F_1,481_=20.7, p< 0.001) compared to non-OUD. Inflammatory markers were also elevated in individuals with OUD, including CRP (F_1,481_=9.4, p=0.002) and ESR (F_1,481_=7.4, p= 0.007), and statistically mediated group differences in AIP and HbA1c, respectively. Our results are consistent with prior evidence of metabolic dysfunctions in individuals with OUD and suggest inflammation as a contributing mechanism. Targeting metabolic health and inflammation may offer new avenues for improving long-term health outcomes in OUD.

## 1. INTRODUCTION

Opioid use disorder (OUD) remains a major public health crisis in the US (Administration, 2024; Ahmad et al., 2024). Among the various health consequences of chronic opioid misuse are metabolic sequelae, including weight gain, inflammation, and sugar liking, but these effects remain relatively understudied and findings are widely inconsistent (Byanyima et al., 2023). A clearer characterizing of the metabolic alterations associated with OUD is important for improving treatment strategies and long-term health outcomes in individuals with OUD.

Several lines of evidence link active opioid use to weight loss. In preclinical rodent models, mice and rats receiving twice-daily injection of escalating does of morphine (Li et al., 2022a; Li et al., 2022b) and rats given unlimited access to heroin self-administration (Chen et al., 2006) consumed less food and exhibited significantly lower body weight compared to saline-treated controls. In concordance with these findings, human observational studies also associated illicit use of opioid within the past 5-years (Tang et al., 2010) and past-year OUD diagnosis (Hu et al., 2018) with lower body mass index (BMI) and lower prevalence of obesity. In qualitive interview with individuals in recovery from substance use disorders, including OUD, participants recalled loss of appetite and rarely eating during active addiction, resulting in severe weight loss (Cowan and Devine, 2008). However, participants reported weight gain during recovery (Cowan and Devine, 2008). Furthermore, while opioid agonists methadone and buprenorphine are highly effective medications for OUD (MOUDs) (Ma et al., 2019; Wakeman et al., 2020), their use have been associated with significant weight gain (Baykara and Alban, 2019; Carr et al., 2023; Manza et al., 2022). In addition to body weight, MOUDs also appear to influence other aspects of metabolic health. For example, patients treated with methadone have higher rates of metabolic syndrome compared to those treated with buprenorphine (Elman et al., 2020), and longer methadone exposure was associated with poorer metabolic profiles (Elman et al., 2020; Vallecillo et al., 2018).

OUD- and treatment-related increases in body weight, specifically visceral fat mass, may contribute to the elevated risk of metabolic dysfunction in OUD (Kawai et al., 2021). White adipose tissue, the predominant fat depot in the body, is an endocrine organ that secretes adipokines regulating various metabolic and inflammatory processes. Increased adiposity facilitates a phenotypic switch in white adipose tissue, promoting a chronic state of low-grade inflammation (Gutierrez et al., 2009; Kawai et al., 2021). As such, several cross-sectional clinical studies have demonstrated correlations between abdominal adiposity and markers of inflammation, including C-reactive protein (CRP) and cumulative inflammation index (Ferrell et al., 2024; Lapice et al., 2009). These findings were corroborated by a genetic study showing that individuals with single-nucleotide polymorphisms in adiposity-related genes, *FTO* and *MC4R*, exhibited higher body mass index (BMI) and CRP levels, suggesting a causal link between adiposity and inflammation (Welsh et al., 2010). Chronic inflammation, in turn, contributes to various forms of metabolic dysfunction. In healthy volunteers, experimentally induced inflammation using low-dose lipopolysaccharide triggered immediate elevations in inflammatory markers (e.g., CRP), altered gene expression related to insulin receptor signaling, and decreased insulin sensitivity within 24 hours (Mehta et al., 2009). Furthermore, inflammation appears to play a role in dyslipidemia. For example, polymorphisms in the *CRP* gene have been linked to susceptibility to dyslipidemia (Wei et al., 2014). Moreover, cross-sectional analyses have positively associated higher CRP levels with increased low-density lipoprotein cholesterol (LDL-C), total cholesterol (Total-C), and triglyceride levels, and with lower high-density lipoprotein cholesterol (HDL-C) levels (Miller et al., 2005).

Metabolic syndrome is characterized by the presence of three or more of the following criteria: increased waist circumference, elevated triglycerides, reduced high-density lipoprotein cholesterol, elevated blood pressure, and elevated glucose. Metabolic syndrome increases the risk for metabolic disorders, including diabetes mellitus and cardiovascular diseases (Eckel et al., 2010). This study leveraged data from a natural history protocol at the National Institutes of Health (NIH) Clinical Center to explore the metabolic health of individuals with a lifetime history of OUD. The primary objective was to compare metabolic and inflammatory parameters between individuals with lifetime OUD and matched non-OUD controls. Given data availability, we used BMI as a proxy for waist circumference, glycated hemoglobin A1c (HbA1c; an index of average blood glucose levels over the past three months) in lieu of fasting glucose levels, and excluded blood pressure measurements. The secondary objective was to examine the role of elevated BMI and inflammation in serially mediating metabolic impairments in OUD. Metabolic impairments were indexed by the atherogenic index of plasma (AIP), a lipid biomarker derived from triglycerides and HDL-C (Zhu et al., 2018), and HbA1c. Lastly, the tertiary objective was to explore the impact of MOUDs and OUD characteristics on metabolic and inflammatory markers. The findings from this study may have implications for guiding treatment strategies to mitigate future metabolic complications in this population.

## 2. METHODS

### 2.1 Participants

The study included participants who completed a natural history protocol at the NIH Clinical Center (Unit and Clinic Evaluation, Screening, Assessment, and Management, ClinicalTrials.gov Identifier: NCT02231840). All participants provided written informed consent to participate in this study as approved by the NIH. The assessments were completed between February 2015 and May 2024. Multivariate outliers were identified using Mahalanobis Distance at a threshold of probability variables of less than 0.001 (Leys et al., 2018). All participants were assessed for substance use disorders and other psychiatric diagnoses using the Structured Clinical Interview for DSM (SCID), version IV or 5, and smoking status with the Fagerström test for nicotine dependence (Heatherton et al., 1991). Individuals with non-OUD had no current or past diagnosis of OUD but most had present or past histories of other SUD, most frequently AUD (70%), and were matched to individuals with a lifetime history of OUD based on race, ethnicity, sex, socioeconomic status (indexed using a 9-tier household income scale), age, and years of education using propensity score matching with a strict match tolerance of 0.1. However, the prevalence of other substance use disorders (alcohol, cannabis, and stimulants; Nagelkerke R^2^= 0.30, p< 0.001) and smoking status (Nagelkerke R^2^= 0.14, p< 0.001) were significantly higher in individuals with OUD than non-OUD, and were included as covariates in subsequent analysis. Out of the original sample of n=1358 participants, the final sample included 527 participants (n=281 OUD and n=246 non-OUD). **Table 1** describes the clinical and demographic characteristics of the participants. Participants eligible for a neuroimaging study (NCT03190954) were invited for a second screening visit, during which MOUDs were assessed in greater details.

**Table 1.**
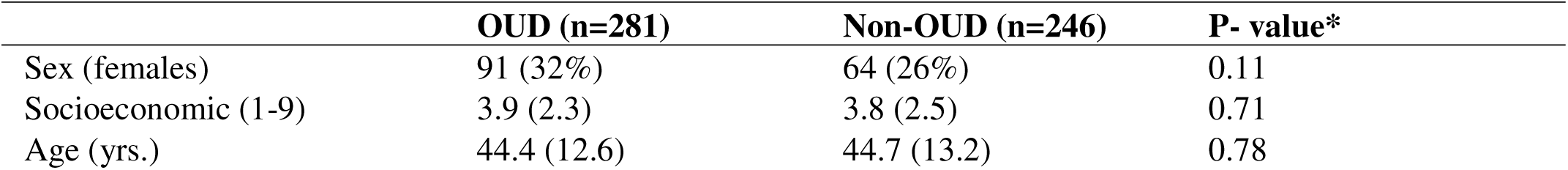
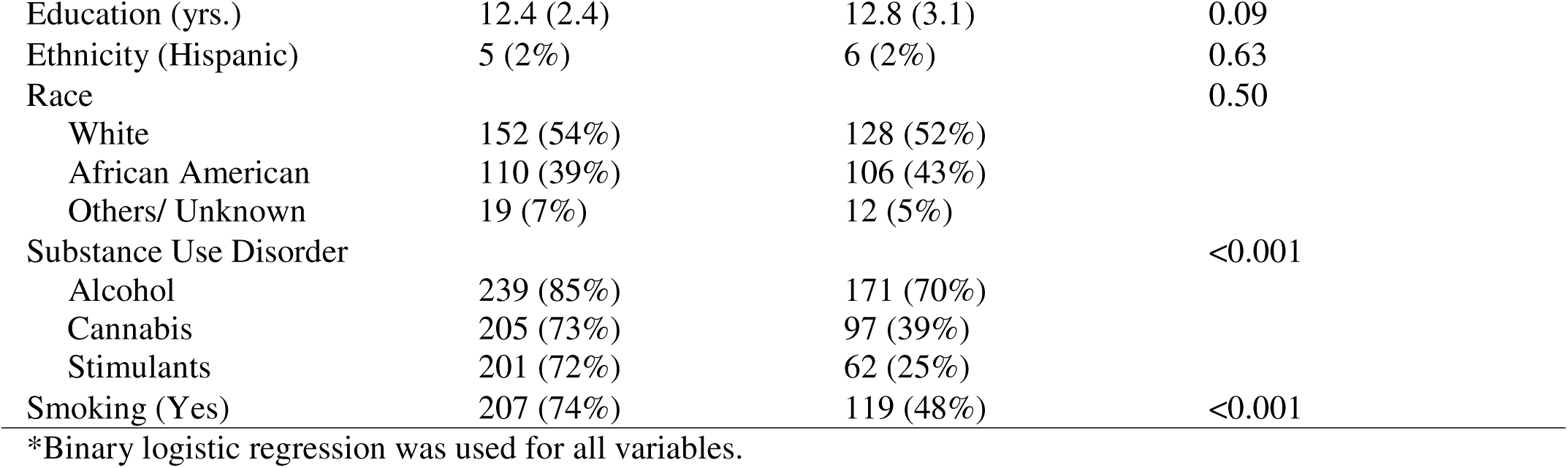
Participant characteristics: number of occurrences (%) or mean (standard deviation)

### 2.2 Data analyses

All analyses were conducted in SPSS (IBM Corp., Armonk, NY, USA). For the primary objective, multivariate analysis of covariance (MANCOVA) was utilized to examine the effects of group (individuals with OUD *vs.* matched non-OUD controls) on BMI, HbA1c, lipid profiles [i.e., total-C, HDL-C, LDL-C, and triglycerides (TG)], atherogenic index of plasma (AIP, calculated as log_10_TG/ HDL) (Zhu et al., 2018), and inflammatory markers [i.e., log_10_ C-reactive protein (CRP) and erythrocyte sedimentation rate (ESR)], while controlling for other substance use disorders (i.e., alcohol, cannabis, and stimulants) and smoking status as covariates. CRP levels were log-transformed due to their skewed distribution. Subsequent univariate analyses examined group differences on individual dependent variables, with Bonferroni correction for multiple comparisons. Partial η^2^ (η ^2^) was used to indicate the effect sizes (small, 0.01; medium, 0.06; large, 0.14).

The secondary objective was to test our hypothesis that OUD-associated impairments in AIP and HbA1c were serially mediated by elevated BMI and inflammation (**Figure 2A**). To this end, we performed serial mediation analysis using PROCESS v.4.2 by Andrew F. Hayes (Model 6) for 4 models consisting of one independent variable (OUD), two serial mediators (BMI → log_10_CRP and BMI → ESR), and two outcomes (AIP and HbA1c), while controlling for other substance use disorders and smoking status as covariates. We utilized a bootstrap of 5000 samples and a 99% confidence interval.

For the tertiary objective of examining the impact of OUD characteristics on metabolic and inflammatory parameters, we conducted a one-way ANOVA between the three medication groups (no medication, methadone, and buprenorphine) in a subset of participants for whom medication data were available (i.e., n=30 on methadone, n=19 on buprenorphine, and n=11 on no medication). Additionally, we performed linear regression analyses using current OUD diagnosis (yes=1, n=190; no=0, n=91) and the number of SCID criteria met for lifetime OUD (n=146) as predictors.

## 3. RESULTS

There was a significant difference between individuals with OUD and non-OUD on the combined set of dependent variables (MANCOVA F_9, 473_= 9.1, p< 0.001, Wilks’ λ= 0.9, η_p_^2^=0.1) (**Figure 1**). Follow-up univariate tests showed that, compared to non-OUD, individuals with OUD had significantly higher BMI (**Figure 1A**, F_1, 481_= 12.9, p< 0.001, η ^2^=0.03), altered lipid profile, including lower HDL-C (**Figure 1C**, F_1, 481_= 46.2, p< 0.001, η_p_^2^=0.09), higher LDL-C (**Figure 1D**, F = 11.9, p< 0.001, η_p_^2^=0.02), and higher AIP (**Figure 1F**, F_1, 481_= 20.7, p< 0.001, η_p_^2^=0.04), and elevated HbA1c (**Figure 1G**, F_1, 481_= 10.5, p=0.001, η_p_^2^=0.02). Moreover, individuals with OUD *vs.* non-OUD had greater levels of inflammatory markers, ESR (**Figure 1H**, F_1, 481_= 7.4, p=0.007, η_p_^2^=0.015) and log_10_CRP (**Figure 1I**, F_1, 481_= 9.4, p= 0.002, η_p_^2^=0.02). No significant differences in total cholesterol (**Figure 1B**, F_1, 481_= 1.6, p=0.2, η_p_^2^=0.003) or triglyceride levels (**Figure 1E**, F_1, 481_= 1.7, p=0.2, η_p_^2^=0.003) was observed.

**Figure 1:**
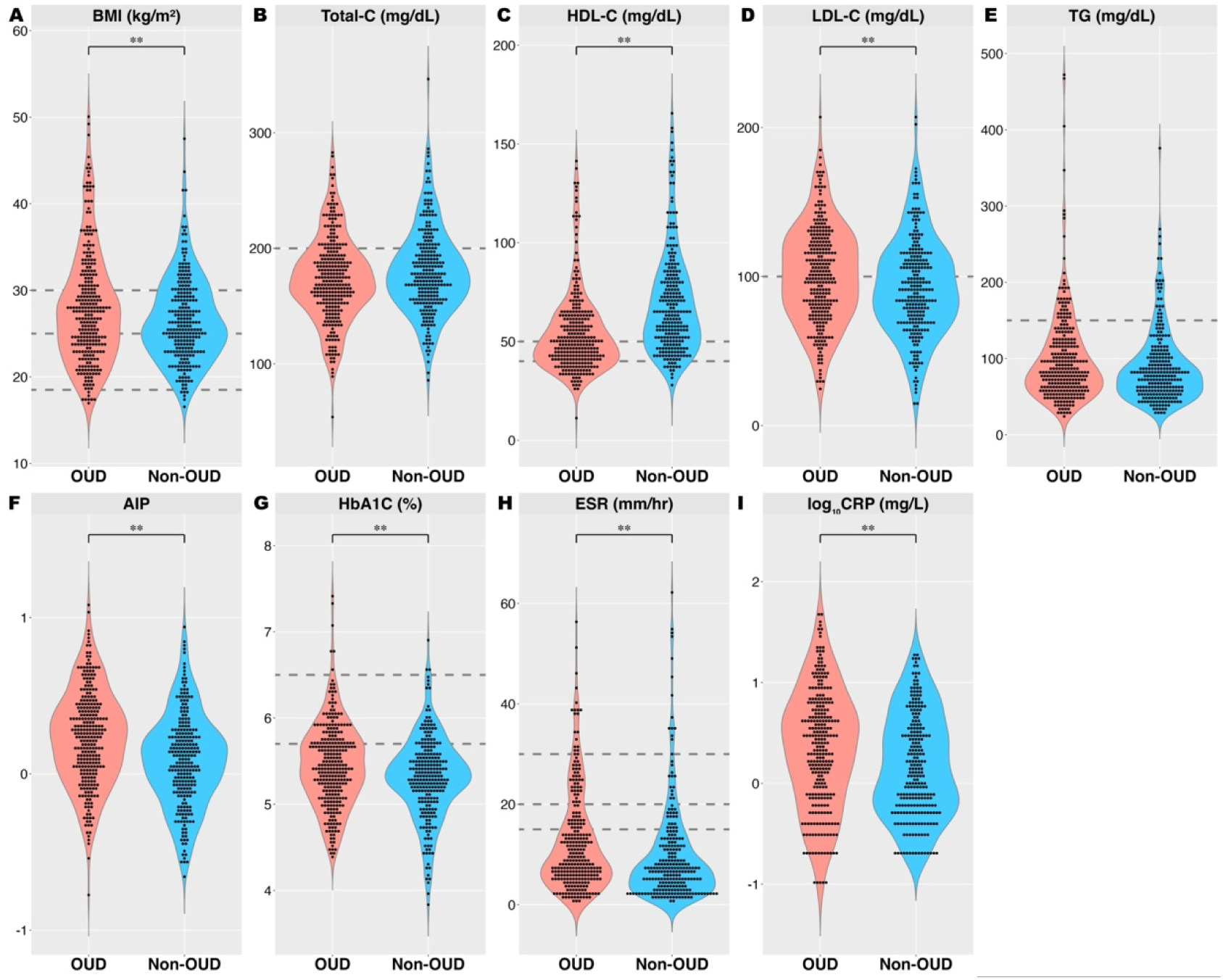
Metabolic and inflammatory parameters in individuals with OUD and matched non-OUD controls. Violin plots of (A) BMI [normal weight 18.5-24.9; overweight 25.0-29.9; obese ≥30 kg/m^2^ (Zierle-Ghosh and Jan, 2025)], (B) Total-C [<200 mg/ dL], (C) HDL [>40 (men) and >50 mg/ dL (women)], (D) LDL [<100 mg/ dL], (E) TG [< 150 mg/ dL] (Pappan et al., 2025), (F) AIP, (G) HbA1c [normal <5.7; prediabetes 5.7-6.4; diabetes ≥ 6.5% (Eyth et al., 2025)], (H) ESR [under 50 years: men ≤15 and women ≤20 mm/ hr.; older than 50 years: men ≤20 and women ≤30 mm/hr. (Tishkowski and Zubair, 2025)], and (I) log-transformed CRP in individuals with OUD and non-OUD. Dotted lines denote the clinical reference range. **Indicate significance p< 0.01. Abbreviations: AIP, atherogenic index of plasma (log_10_TG/HDL); BMI, body mass index, CRP, C-reactive protein; ESR, erythrocyte sedimentation rate; HbA1c, hemoglobin A1c; HDL-C, high-density lipoprotein cholesterol; LDL-C, low-density lipoprotein cholesterol; TG, triglycerides; Total-C, total cholesterol; OUD, opioid use disorder.

The mediation model examining the role of BMI→ log_10_CRP in serially mediating the effects of OUD on AIP and HbA1c demonstrated significant direct (AIP: Effect= 0.11, 99% CI [0.03, 0.2]; HbA1c: Effect = 0.15, 99% CI [0.04, 0.3]) and total indirect (AIP: Effect = 0.04, 99% CI [0.02, 0.06]; HbA1c: Effect = 0.029, 99% CI [0.003, 0.06]) effects. The relative indirect effects of BMI→ log_10_CRP were significant for AIP (Effect = 0.010, 99% CI [0.002, 0.02]), but not for HbA1c (**Table 2** and **Figure 2A**). Analyses with mediators BMI→ ESR indicated significant direct and total indirect effects of OUD on AIP (direct: Effect = 0.11, 99% CI [0.03, 0.2]; total indirect: Effect = 0.038, 95% CI [0.01, 0.07]) and HbA1c (direct: Effect = 0.12, 99% CI [-0.02, 0.3]; total indirect: Effect = 0.041, 99% CI [0.006, 0.09]). The relative indirect effects of BMI→ ESR in serially mediating the relationship between OUD and HbA1c (Effect = 0.007, 95% CI [0.001, 0.02]) were significant, but were not significant for AIP (**Table 2** and **Figure 2B**).

**Table 2:**
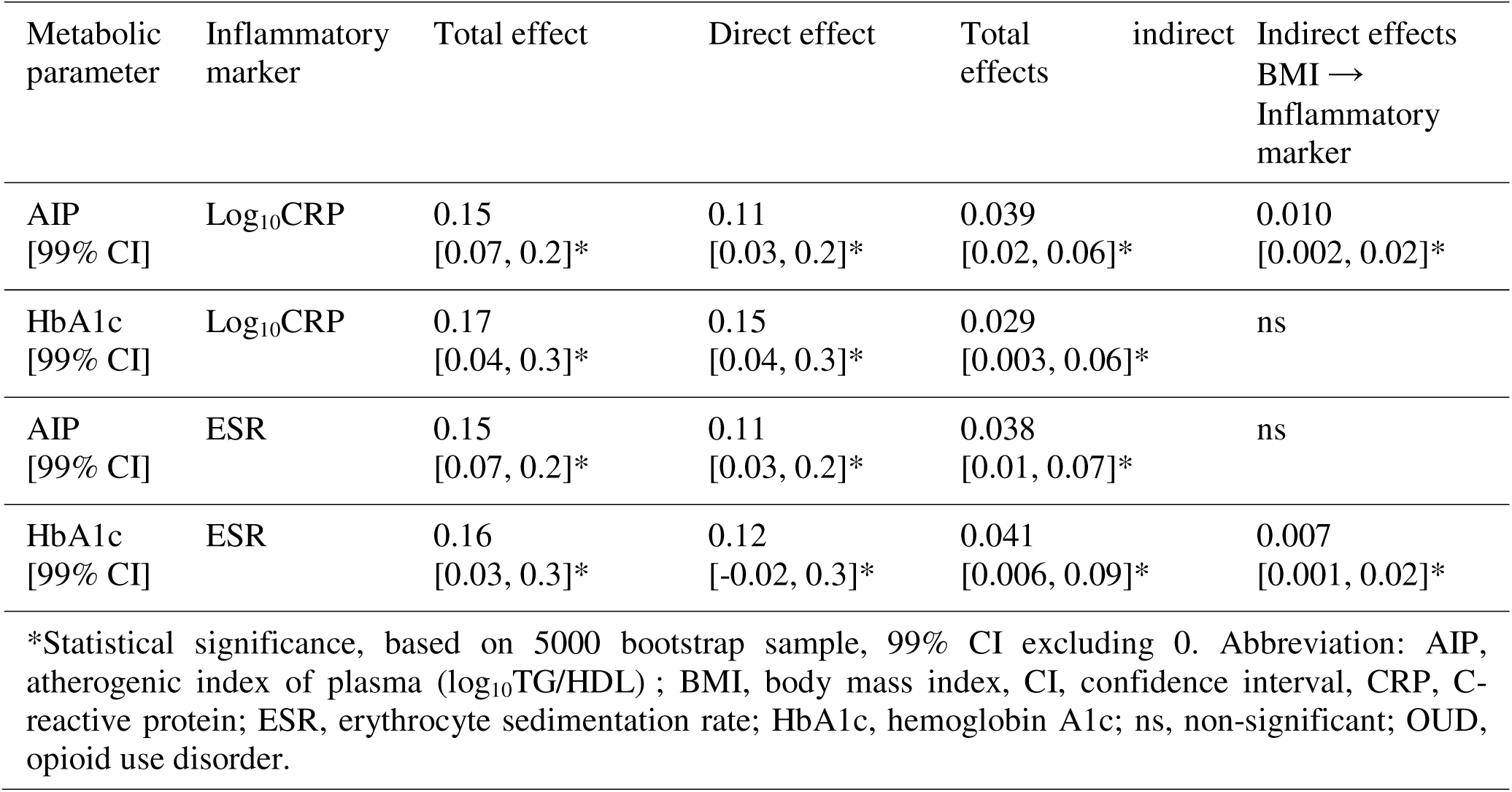
Total and direct effects of OUD on metabolic parameters and indirect effects serially mediated through BMI. ➔ **inflammation.**

**Figure 2:**
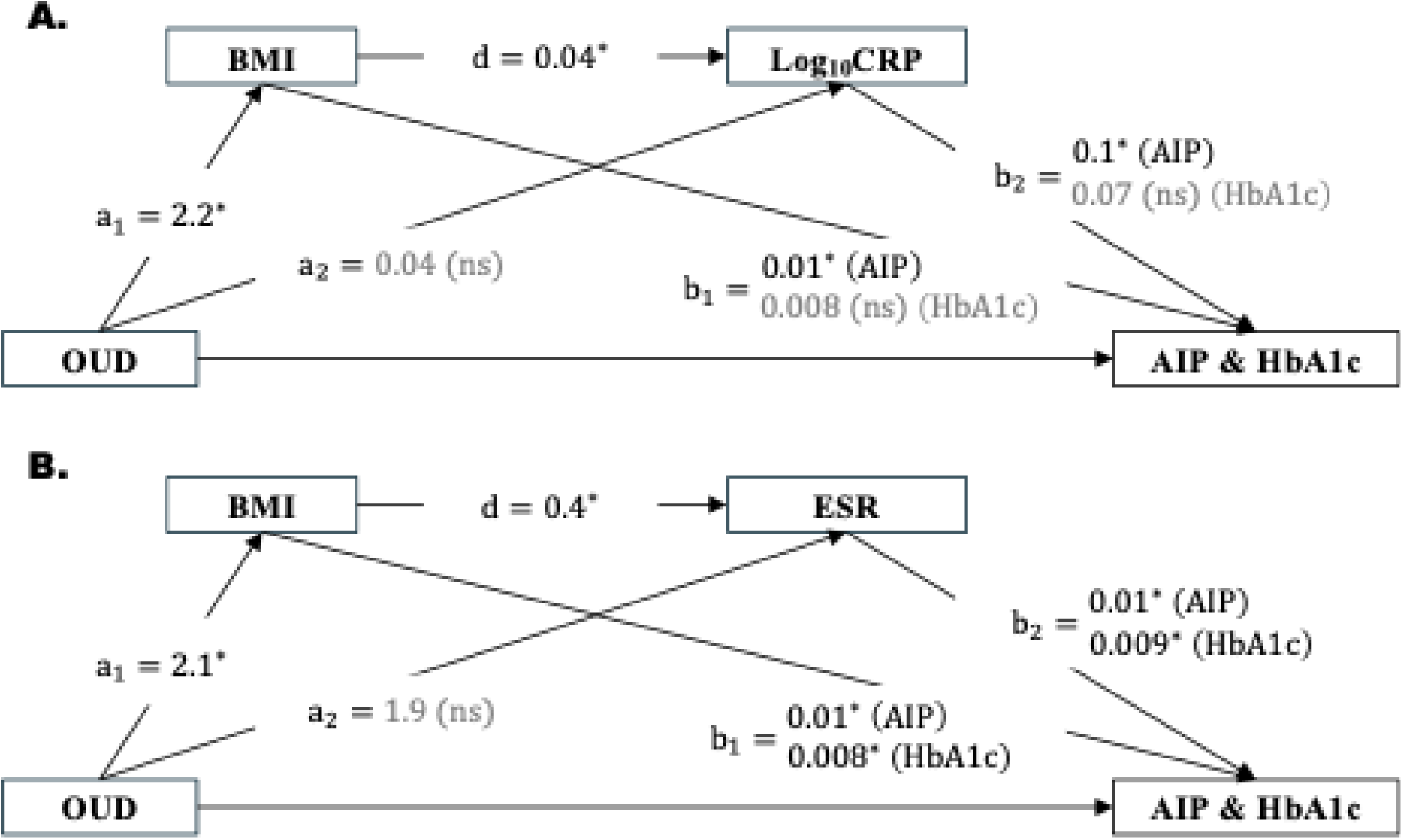
Serial mediation model. We hypothesized that OUD-associated impairment in AIP and HbA1c were serially mediated by elevated BMI and inflammatory markers, (A) log_10_CRP and (B) ESR. Regression coefficients are presented. *, p< 0.05; ns, non-significant. Abbreviations: AIP, atherogenic index of plasma (log_10_TG/HDL) ; BMI, body mass index, CRP, C-reactive protein; ESR, erythrocyte sedimentation rate; HbA1c, hemoglobin A1c; OUD, opioid use disorder.

In individuals with a lifetime diagnosis of OUD, HbA1c differed across groups (F_2, 59_= 4.3, p=0.018) and *post hoc* analysis revealed lower levels in individuals on no medications compared to those on buprenorphine (p< 0.05, **Figure 3G**). ESR also demonstrated significant group effects (**Figure 3H**, F_2, 57_= 5.2, p=0.008), with lower levels observed in individuals on buprenorphine than on methadone (p <0.05). There were no significant effects of medication status (recovery with no medications *vs*. methadone *vs.* buprenorphine) on BMI (**Figure 3A**), lipids, including total-, HDL-, and LDL-cholesterol, triglycerides, and AIP (**Figures 3B-F**) and log_10_CRP (**Figure 3I**, p>0.05). Individuals included *vs.* not included in the sub-analysis did not significantly differ in any demographic variables or prevalence of lifetime diagnosis of other substance use disorders (**Supplemental Table 1S**). We note that this analysis was conducted in a much smaller subsample of participants and was therefore not powered to detect effect sizes comparable to the primary analyses, which may limit the generalizability of the findings to a broader population of individuals with OUD. In addition, there was a significant group difference in the proportion of individuals with a current OUD diagnosis (within the past 12 months), which may have confounded the results (demographics provided in **Supplemental Table 2S**). Indeed, a diagnosis of current OUD, at least within the past 12 months, was associated with higher HbA1c (F_1, 277_= 38.3, p< 0.001, R^2^= 0.12), log_10_CRP (F_1, 278_= 18.0, p< 0.001, R^2^= 0.06), and ESR (F_1, 272_= 20.5, p< 0.001, R^2^= 0.07), but not with BMI or AIP (p>0.05). Results of t-test comparisons between individuals with and without current OUD diagnosis are presented in **Supplemental Figure 4S**. The number of SCID criteria met for the diagnosis of lifetime OUD did not correlate with any metabolic or inflammatory markers (p > 0.05) (**Table 3**).

**Figure 3:**
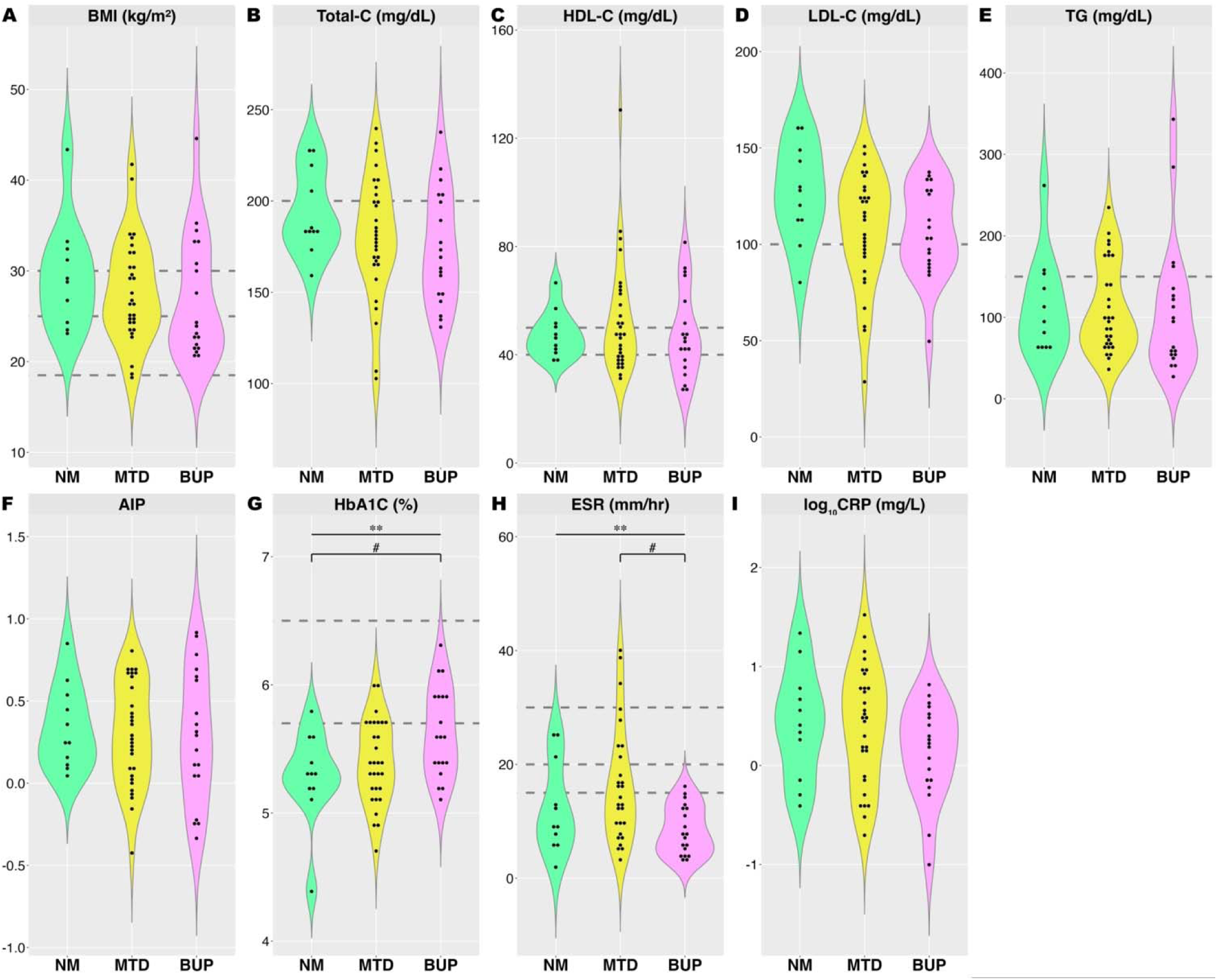
Metabolic and inflammatory parameters in individuals with OUD by medication. Violin plots of (A) BMI, (B) Total-C, (C) HDL, (D) LDL, (E) TG, (F) AIP, (G) HbA1c, (H) ESR, and (I) log-transformed CRP in individuals with OUD on no medication (n=11), methadone (n=30), or buprenorphine (n=19). Dotted lines denotes the clinical reference range. **One-way ANOVA analysis indicates significant main effect of group, p< 0.05. #Indicates differences between two groups. Abbreviations: AIP, atherogenic index of plasma (log_10_TG/HDL); BMI, body mass index; BUP, buprenorphine; CRP, C-reactive protein; ESR, erythrocyte sedimentation rate; HbA1c, hemoglobin A1c; HDL-C, high-density lipoprotein cholesterol; LDL-C, low-density lipoprotein cholesterol; MTD: methadone; NM: no medication; TG, triglycerides; Total-C, total cholesterol; OUD, opioid use disorder.

**Table 3:**
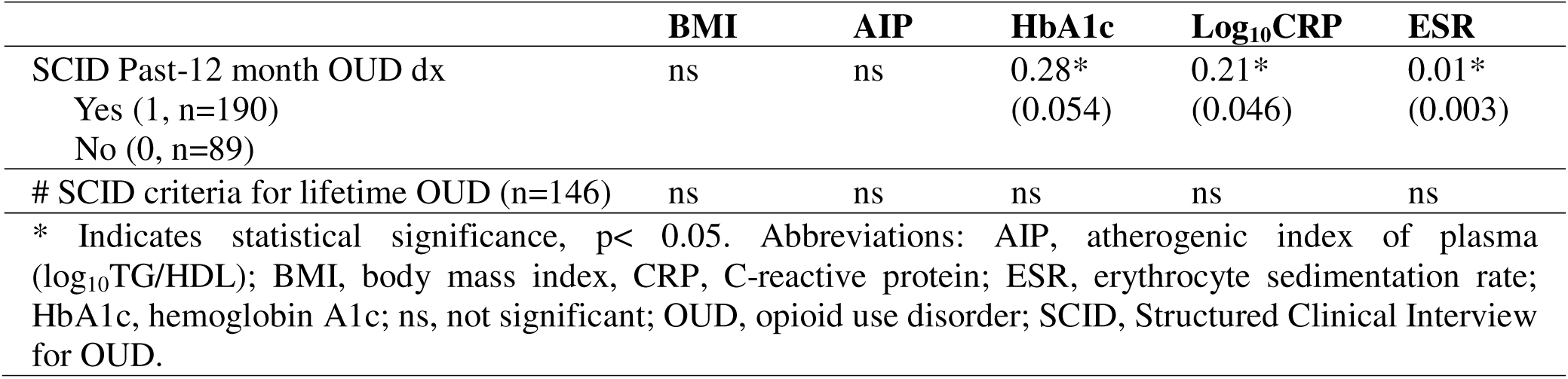
Linear regression of OUD characteristics with metabolic and inflammatory markers. Reported as unstandardized coefficient B (standard error).

## 4. DISCUSSION

Our study emphasized the importance of incorporating biopsychosocial considerations in the treatment of individuals with OUD. Prior research has reported that higher BMI is positively associated with increased likelihood of naltrexone treatment adherence in human patients with OUD (Li et al., 2023) and that diet-induced obesity attenuates neural responses to opioid withdrawal in rodents (Li et al., 2022b), suggesting a potential beneficial effect of elevated body weight on initial treatment outcomes. However, this study identified several metabolic impairments in individuals with OUD, including elevated BMI, altered lipid profiles, and higher HbA1c levels, compared to matched non-OUD controls. The average BMI of both groups falls within the overweight range (Zierle-Ghosh and Jan, 2025), with higher values observed in individuals with OUD (28.0 ± 5.9 kg/m^2^) *vs.* non-OUD (26.5 ± 4.9 kg/m^2^), raising additional clinical concerns. Although mean lipid, HbA1c, and ESR values were within normal ranges, poorer values, even within the normal range, may progressively increase the risks for developing metabolic disorders later in life (Bonora et al., 2011; Leiherer et al., 2021). In 1990, the American Dietetic Association published a position paper acknowledging substance use-induced physical adaptations and malnutrition and proposing the inclusion of registered dieticians in the rehabilitation team (1990). Despite this, a survey of substance use treatment facilities in Los Angeles revealed that only 30.5% of the facilities offered some form of on-site nutritional counseling, and more alarmingly, only 6.3% of the facilities utilized registered dietitians (Wiss et al., 2019). Our findings of poorer metabolic health in individuals with OUD further emphasize the need for incorporating nutritional intervention by providers for substance use disorder treatments, as previously recommended, including nutrition screening and assessments, nutritional counseling/ education on nutrition life skills, incorporation of healthier meal choices in residential treatment programs, and nutritional supplement interventions (1990; Chavez and Rigg, 2020).

Previous studies have linked active opioid use to lower body weight and reduced fat mass (Hu et al., 2018; Nazrul Islam et al., 2002; Tang et al., 2010), (Byanyima et al., 2023). Weight loss during active use may be attributable to opioid-induced neuroadaptations in reward and reinforcement pathways that prioritize drug seeking at the expense of other goal-directed behaviors, including eating (Chavez and Rigg, 2020). Individuals with OUD often report allocating limited financial resources to purchasing opioids in lieu of food, skipping meals, and reduced appetite (Chavez and Rigg, 2020; Mahboub et al., 2021). In contrast, during treatment and recovery, individuals often report increased appetite and food consumption (Cowan and Devine, 2008; Neale et al., 2012). Although the initial weight gain may be perceived as compensation for prior weight loss, sustained increases over time may contribute to the development of overweight and obesity (Cowan and Devine, 2008; Gottfredson and Sokol, 2019; Mahboub et al., 2021). Additionally, a growing body of evidence indicates that opioid agonist MOUD therapies (methadone and buprenorphine) may independently contribute to weight gain (Baykara and Alban, 2019; Carr et al., 2023; Fenn et al., 2015; Manza et al., 2022). Our finding of higher BMI among individuals with OUD is consistent with patterns observed in those recovering from OUD and receiving MOUD, but contrasts with findings in individuals who are actively using illicit opioids or have untreated OUD. However, further contextualization of these findings within the existing literature was limited by the lack of available data addressing the potential confounding effects of active illicit opioid use, opioid withdrawal, and treatment with full or partial opioid agonist-based MOUD, and therefore further research is warranted.

Several mutually non-exclusive hypotheses have been put forth in attempts to explain weight gain in individuals with OUD during recovery and MOUD treatment. Neurophysiology underlying compulsive reward seeking in substance use disorders and compulsive food intake may overlap, placing individuals with OUD at an increased risk for comorbid eating disorders (i.e., binge eating and food addiction) (Canan et al., 2017). According to the *Propensity for Behavioral Addiction Hypothesis*, in individuals susceptible to compulsive eating, active opioid use suppresses appetite and masks these tendencies, but during recovery/detoxification, the absence of opioids can lead to a resurgence of maladaptive eating patterns and subsequent weight gain (Gottfredson and Sokol, 2019). Additionally, commonly prescribed MOUDs, opioid agonists methadone and buprenorphine, may further contribute to weight gain, as opioids have been implicated in the hedonic/pleasurable aspects of eating (Gosnell and Levine, 2009). Studies assessing dietary patterns in individuals with OUD receiving MOUD report greater preference for energy-dense, nutritionally empty, palatable foods, such as simple carbohydrates (e.g., bread), sweets, and “junk” food and reduced consumption of fruits and vegetables (Kheradmand and Kheradmand, 2020; Nolan and Scagnelli, 2007; Ochalek et al., 2021; Peles et al., 2016). Reflecting on periods of active use, individuals with OUD similarly report a preference for sweet foods and reliance on quick, convenient options (e.g., sweets, tinned foods, breakfast cereal) (Neale et al., 2012). However, caloric intake from these energy-dense food may not compensate for overall reduction in food consumption, as meal skipping, prioritization of opioid use over eating, and suppressed appetite, potentially driven by opioid-related alteration in dopaminergic reward and reduced sensitivity to natural rewards, may collectively limit energy intake and attenuate weight gain (Chavez and Rigg, 2020). Thirdly, social and psychological factors in individuals with OUD in recovery, such as eating alone, fear of meeting encountering individuals under the influence of substances of abuse at free food distribution centers, and emotional stress associated with in maintaining sobriety, may further decrease motivation to exert the effort to prepare healthy meals and limit access to nutritious foods (Furulund et al., 2024; Mahboub et al., 2025).

In this study, we found individuals with OUD had altered lipid profile, including lower HDL-C, higher LDL-C, and higher AIP, and elevated HbA1c., compared to those without OUD. Prior research examining the effects of opioids on metabolic health have yielded mixed results, reviewed in (Byanyima et al., 2023). Studies have associated opioid use with lower risk for metabolic syndrome (Bagheri-Hosseinabadi et al., 2022), lower cholesterol, including the “good” cholesterol HDL-C (Kazemi et al., 2021), and impaired glucose tolerance (Karam et al., 2004). These inconsistencies may reflect heterogeneity in study populations and exposure types, including occasionally using naturally derived opium (e.g., crude opium or opium sap), dependent on synthetic opioids (e.g. heroin), differences between past exposure *vs.* active use, and studies with unspecified opioid exposures. Nonetheless, studies of individuals with OUD receiving opioid agonist treatment, particularly methadone, have more consistently demonstrated poorer metabolic parameters (Elman et al., 2020; Fareed et al., 2009; Vallecillo et al., 2018). Given that increased adiposity and associated chronic low-grade inflammation contribute to metabolic dysfunction, we hypothesized that elevated BMI and inflammatory markers would significantly and serially mediate the association between OUD and both elevated AIP, indicative of dyslipidemia, and elevated HbA1c, a marker of glycemic control. Although significant serial mediation by BMI and inflammatory markers was observed, significant direct effects of OUD on these metabolic outcomes persisted after accounting for these factors, suggesting additional contributory factors that warrant further investigation.

In a subset of individuals with OUD for whom data were available, we explored the correlation between OUD characteristics and biomarkers of metabolic and inflammatory status. Contrary to previous studies linking full opioid agonist methadone but not partial agonist buprenorphine to poorer metabolic markers (Elman et al., 2020; Fareed et al., 2009; Vallecillo et al., 2018), we did not observe significant associations between methadone treatment and metabolic markers (i.e., BMI, AIP, and HbA1c) but instead found buprenorphine treatment to be associated with higher HbA1c. This finding, coupled with our observation that methadone use correlated positively and buprenorphine use negatively with ESR, contradicts our hypothesis and results from serial mediation analyses that elevated BMI and inflammatory status contribute to impairments in HbA1c. There were group differences in smoking status and in the proportion of individuals with a current (within the past 12 months) diagnosis of OUD (no medication: 27%, methadone: 83%, buprenorphine: 95%), which may have confounded our findings (**Supplemental Table 1S**). Furthermore, these analyses were conducted in a smaller subsample (n=30 for methadone, n=19 for buprenorphine, and n=11 for no medication), increasing the likelihood of random error and limiting the generalizability to a broader population of individuals with OUD. These limitations highlight the need for future research on the complex interrelationship between MOUDs, inflammation, and metabolic health. Additionally, our findings indicated that higher inflammation and HbA1c were associated with having a current (within the past 12 months) OUD diagnosis (as per SCID), but not with OUD severity, defined by the number of SCID diagnostic criteria met. Therefore, sustained recovery from OUD may have the potential to ameliorate some of the inflammatory and metabolic impairments, although further investigation is needed.

Our study included a relatively large sample size of individuals with OUD who were matched to non-OUD controls based on race, ethnicity, sex, socioeconomic status, age, and years of education. These variables are established determinants of metabolic health (Blanquet et al., 2019; Moore et al., 2017), matching for which minimized their potentially confounding effects, strengthening the likelihood that the observed group differences in metabolic and inflammatory markers were attributable primarily to OUD. Nonetheless, our study had several limitations. Firstly, the correlative nature of our analyses precludes the establishment of causal relationships, and unknown confounders may have influenced our study findings. In the Supplemental Material, we tested and discussed an alternative mechanism, in which elevated BMI and metabolic dysfunction serially mediated the relationship between OUD and heightened inflammatory state (**Supplemental Figure 2S** and **Table 2S**). Secondly, we utilized a convenient sample of individuals with OUD and non-OUD, most of whom had a history of AUD of diverse severity. Because metabolic and inflammatory markers were not the primary objective of the study, we were limited by data availability. As such, no blood pressure or waist circumference measurements were collected, precluding us from diagnosing metabolic syndrome and assessing the impact of OUD on its prevalence. It is also important to note that although waist circumference and BMI are highly correlated, the use of BMI as a proxy for central adiposity may limit the precision of our assessment (Ross et al., 2020). Moreover, the study was limited by the availability of inflammatory markers collected in the individuals, and future studies using other markers, including cytokines (e.g., interleukins and tumor necrosis factor), may provide additional insights.

In our study population, a concurrent AUD diagnosis was present in a greater proportion of individuals with OUD (85%) than in non-OUD (70%, χ^2^_1_= 18.34, p< 0.001). Extensive studies have examined the metabolic consequences of alcohol use and suggested a J-shaped relationship, whereby low-to-moderate consumption was associated with improved lipid levels (Brien et al., 2011; Suzuki et al., 2025) and reduced risk for diabetes (Baliunas et al., 2009). However, concerns have been raised about the observational nature of these studies and that the findings may be influenced by underlying health status, age, and other confounding factors (Fillmore et al., 2007). In contrast, chronic heavy alcohol use in individuals with AUD has been associated with adverse metabolic outcomes, including impaired lipid levels (Rosoff et al., 2019; Wiers et al., 2021) and increased risk for diabetes (Baliunas et al., 2009), and a prevalence of metabolic syndrome ranging from approximately 13.2% (Hernández-Rubio et al., 2022) to 21.8% (Vancampfort et al., 2016). AUD was included as a covariate in our analysis to mitigate its potential confounding effects, but its comorbidity with OUD and its known metabolic impact may exert residual effects on the interpretation of our findings. In the Supplemental analysis, we compared metabolic and inflammatory markers in control individuals and individuals with OUD only, AUD only, and OUD+AUD, and found greater metabolic impairments in individuals with OUD than AUD (**Supplemental Figure 3S**). Therefore, future research is needed to disentangle the independent and combined effects of OUD and AUD on metabolic risks, thereby developing more targeted interventions to improve health outcomes.

In sum, we found metabolic impairments in individuals with OUD compared to matched non-OUD controls. These impairments, reflected by elevated AIP and HbA1c, were serially mediated by increases in BMI and inflammatory markers. The study findings provide a theoretical framework for understanding metabolic disturbances in OUD and underscore the importance of addressing long-term metabolic health in the treatment of individuals with OUD.

## 5. Funding source

This work was supported by the intramural support from the NIH-National Institute on Alcohol Abuse and Alcoholism Y1AA-3009 (Volkow) and the Division of Intramural Clinical and Biological Research, NIAAA, including the 1SE Inpatient Behavioral Health Unit and the 1SE Outpatient Clinic (Diazgranados). Dr Li was supported by the National Institutes of Health (NIH) grant AA031746, Dr. Wiers by AA031570, AA031088, AA031337, and Dr. Shi by DA051709 and the NARSAD Young Investigator Grants from the Brain and Behavior Research Foundation #30780.

## Data Availability

All data produced in the present study are available upon reasonable request to the authors

## Notes

### Competing Interest Statement

The authors have declared no competing interest.

### Clinical Trial

NCT02231840

### Author Declarations

The study included participants who completed a natural history protocol at the NIH Clinical Center (Unit and Clinic Evaluation, Screening, Assessment, and Management; ClinicalTrials.gov Identifier: NCT02231840; NIAAA protocol 14-AA-0181). The protocol was reviewed and approved by the National Institutes of Health Institutional Review Board (NIH IRB), which serves as the ethics committee for the NIH Intramural Research Program. Ethical approval was granted prior to study initiation (January 21, 2015). All participants provided written informed consent to participate in this study.

## References

1. 1990. Position of the American Dietetic Association: nutrition intervention in treatment and recovery from chemical dependency. Journal of the American Dietetic Association 90, 1274+.

2. Administration, S.A.a.M.H.S., 2024. 2023 Companion infographic report: Results from the 2021, 2022, and 2023 National Surveys on Drug Use and Health. Substance Abuse and Mental Health Services Administration.

3. Ahmad, F.B., Cisewski, J.A., Rossen, L.M., Sutton, P., 2024. Provisional drug overdose death counts. National Center for Health Statistics.

4. Bagheri-Hosseinabadi, Z., Khalili, P., Hakimi, H., Jalali, N., Abbasifard, M., 2022. Evaluation of the relationship between opioid addiction and metabolic syndrome and its components in the adult population from Rafsanjan city; a cohort study. Inflammopharmacology 30(6), 2107–2116.

5. Baliunas, D.O., Taylor, B.J., Irving, H., Roerecke, M., Patra, J., Mohapatra, S., Rehm, J., 2009. Alcohol as a Risk Factor for Type 2 Diabetes: A systematic review and meta-analysis. Diabetes Care 32(11), 2123–2132.

6. Baykara, S., Alban, K., 2019. The effects of buprenorphine/naloxone maintenance treatment on sexual dysfunction, sleep and weight in opioid use disorder patients. Psychiatry Research 272, 450–453.

7. Blanquet, M., Legrand, A., Pélissier, A., Mourgues, C., 2019. Socio-economics status and metabolic syndrome: A meta-analysis. Diabetes & Metabolic Syndrome: Clinical Research & Reviews 13(3), 1805–1812.

8. Bonora, E., Kiechl, S., Mayr, A., Zoppini, G., Targher, G., Bonadonna, R.C., Willeit, J., 2011. High-normal HbA1c is a strong predictor of type 2 diabetes in the general population. Diabetes Care 34(4), 1038–1040.

9. Brien, S.E., Ronksley, P.E., Turner, B.J., Mukamal, K.J., Ghali, W.A., 2011. Effect of alcohol consumption on biological markers associated with risk of coronary heart disease: systematic review and meta-analysis of interventional studies. Bmj 342, d636.

10. Byanyima, J.I., Li, X., Vesslee, S.A., Kranzler, H.R., Shi, Z., Wiers, C.E., 2023. Metabolic profiles associated with opioid use and opioid use disorder: a narrative review of the literature. Curr Addict Rep 10(3), 581–593.

11. Canan, F., Karaca, S., Sogucak, S., Gecici, O., Kuloglu, M., 2017. Eating disorders and food addiction in men with heroin use disorder: a controlled study. Eating and Weight Disorders - Studies on Anorexia, Bulimia and Obesity 22(2), 249–257.

12. Carr, M.M., Lou, R., Macdonald-Gagnon, G., Peltier, M.R., Funaro, M.C., Martino, S., Masheb, R.M., 2023. Weight change among patients engaged in medication treatment for opioid use disorder: a scoping review. Am J Drug Alcohol Abuse 49(5), 551–565.

13. Chavez, M.N., Rigg, K.K., 2020. Nutritional implications of opioid use disorder: A guide for drug treatment providers. Psychol Addict Behav 34(6), 699–707.

14. Chen, S.A., O’Dell, L.E., Hoefer, M.E., Greenwell, T.N., Zorrilla, E.P., Koob, G.F., 2006. Unlimited Access to Heroin Self-Administration: Independent Motivational Markers of Opiate Dependence. Neuropsychopharmacology 31(12), 2692–2707.

15. Cowan, J., Devine, C., 2008. Food, eating, and weight concerns of men in recovery from substance addiction. Appetite 50(1), 33–42.

16. Eckel, R.H., Alberti, K., Grundy, S.M., Zimmet, P.Z., 2010. The metabolic syndrome. The Lancet 375(9710), 181–183.

17. Elman, I., Howard, M., Borodovsky, J.T., Mysels, D., Rott, D., Borsook, D., Albanese, M., 2020. Metabolic and Addiction Indices in Patients on Opioid Agonist Medication-Assisted Treatment: A Comparison of Buprenorphine and Methadone. Scientific Reports 10(1), 5617.

18. Eyth, E., Zubair, M., Naik, R., 2025. Hemoglobin A1C, StatPearls. StatPearls Publishing Copyright © 2025, StatPearls Publishing LLC., Treasure Island (FL).

19. Fareed, A., Casarella, J., Amar, R., Vayalapalli, S., Drexler, K., 2009. Benefits of retention in methadone maintenance and chronic medical conditions as risk factors for premature death among older heroin addicts. J Psychiatr Pract 15(3), 227–234.

20. Fenn, J.M., Laurent, J.S., Sigmon, S.C., 2015. Increases in body mass index following initiation of methadone treatment. J Subst Abuse Treat 51, 59–63.

21. Ferrell, K., Brown, I., Amare, A., McNeel, T.S., Ferrell, K., Brown, I., Amare, A., McNeel, T.S., Buckman, D., Jackson, S.H., 2024. Positive association between adiposity and inflammation in US adults: A cross sectional study. Clinical obesity. 14(1).

22. Fillmore, K.M., Stockwell, T., Chikritzhs, T., Bostrom, A., Kerr, W., 2007. Moderate alcohol use and reduced mortality risk: systematic error in prospective studies and new hypotheses. Ann Epidemiol 17(5 Suppl), S16–23.

23. Furulund, E., Druckrey-Fiskaaen, K.T., Carlsen, S.-E.L., Madebo, T., Fadnes, L.T., Lid, T.G., 2024. Healthy eating among people on opioid agonist therapy: a qualitative study of patients’ experiences and perspectives. BMC Nutrition 10(1), 70.

24. Gosnell, B.A., Levine, A.S., 2009. Reward systems and food intake: role of opioids. International Journal of Obesity 33(2), S54–S58.

25. Gottfredson, N.C., Sokol, R.L., 2019. Explaining Excessive Weight Gain during Early Recovery from Addiction. Subst Use Misuse 54(5), 769–778.

26. Gutierrez, D.A., Puglisi, M.J., Hasty, A.H., 2009. Impact of increased adipose tissue mass on inflammation, insulin resistance, and dyslipidemia. Curr Diab Rep 9(1), 26–32.

27. Heatherton, T.F., Kozlowski, L.T., Frecker, R.C., Fagerström, K.O., 1991. The Fagerström Test for Nicotine Dependence: a revision of the Fagerström Tolerance Questionnaire. Br J Addict 86(9), 1119–1127.

28. Hernández-Rubio, A., Sanvisens, A., Bolao, F., Cachón-Suárez, I., Garcia-Martín, C., Short, A., Bataller, R., Muga, R., 2022. Prevalence and associations of metabolic syndrome in patients with alcohol use disorder. Scientific Reports 12(1), 2625.

29. Hu, L., Matthews, A., Shmueli-Blumberg, D., Killeen, T.K., Tai, B., VanVeldhuisen, P., 2018. Prevalence of obesity for opioid- and stimulant-dependent participants in substance use treatment clinical trials. Drug and Alcohol Dependence 190, 255–262.

30. Karam, G.A., Reisi, M., Kaseb, A.A., Khaksari, M., Mohammadi, A., Mahmoodi, M., 2004. Effects of opium addiction on some serum factors in addicts with non-insulin-dependent diabetes mellitus. Addict Biol 9(1), 53–58.

31. Kawai, T., Autieri, M.V., Scalia, R., 2021. Adipose tissue inflammation and metabolic dysfunction in obesity. Am J Physiol Cell Physiol 320(3), C375–c391.

32. Kazemi, M., Bazyar, M., Naghizadeh, M.M., Dehghan, A., Rahimabadi, M.S., Chijan, M.R., Bijani, M., Zahmatkeshan, M., Ghaemi, A., Samimi, N., Homayounfar, R., Farjam, M., 2021. Lipid profile dysregulation in opium users based on Fasa PERSIAN cohort study results. Scientific Reports 11(1), 12058.

33. Kheradmand, A., Kheradmand, A., 2020. Nutritional status in patients under methadone maintenance treatment. Journal of Substance Use 25(2), 173–176.

34. Lapice, E., Maione, S., Patti, L., Cipriano, P., Rivellese, A.A., Riccardi, G., Vaccaro, O., 2009. Abdominal adiposity is associated with elevated C-reactive protein independent of BMI in healthy nonobese people. Diabetes Care 32(9), 1734–1736.

35. Leiherer, A., Ulmer, H., Muendlein, A., Saely, C.H., Vonbank, A., Fraunberger, P., Foeger, B., Brandtner, E.M., Brozek, W., Nagel, G., Zitt, E., Drexel, H., Concin, H., 2021. Value of total cholesterol readings earlier versus later in life to predict cardiovascular risk. eBioMedicine 67.

36. Leys, C., Klein, O., Dominicy, Y., Ley, C., 2018. Detecting multivariate outliers: Use a robust variant of the Mahalanobis distance. Journal of Experimental Social Psychology 74, 150–156.

37. Li, X., Kshatriya, D., Bello, N.T., 2022a. Weight-gain propensity and morphine withdrawal alters locomotor behavior and regional norepinephrine-related gene expression in male and female mice. Pharmacol Biochem Behav 213, 173329.

38. Li, X., Langleben, D.D., Lynch, K.G., Wang, G.J., Elman, I., Wiers, C.E., Shi, Z., 2023. Association between body mass index and treatment completion in extended-release naltrexone-treated patients with opioid dependence. Front Psychiatry 14, 1247961.

39. Li, X., Yeh, C.Y., Bello, N.T., 2022b. High-fat diet attenuates morphine withdrawal effects on sensory-evoked locus coeruleus norepinephrine neural activity in male obese rats. Nutr Neurosci 25(11), 2369–2378.

40. Ma, J., Bao, Y.-P., Wang, R.-J., Su, M.-F., Liu, M.-X., Li, J.-Q., Degenhardt, L., Farrell, M., Blow, F.C., Ilgen, M., Shi, J., Lu, L., 2019. Effects of medication-assisted treatment on mortality among opioids users: a systematic review and meta-analysis. Molecular Psychiatry 24(12), 1868–1883.

41. Mahboub, N., Al Slaybe, M., Mikhael, E., Tohme Khalaf, P., Baltagi, T.K., de Vries, N., Rizk, R., 2025. Dietary and lifestyle adjustments among people who use drugs in opioid substitution treatment and recommendations for program development: A qualitative study. Social Sciences & Humanities Open 12, 102309.

42. Mahboub, N., Rizk, R., Karavetian, M., de Vries, N., 2021. Nutritional status and eating habits of people who use drugs and/or are undergoing treatment for recovery: a narrative review. Nutr Rev 79(6), 627–635.

43. Manza, P., Kroll, D., McPherson, K.L., Johnson, A., Dennis, E., Hu, L., Tai, B., Volkow, N.D., 2022. Sex differences in weight gain during medication-based treatment for opioid use disorder: A meta-analysis and retrospective analysis of clinical trial data. Drug and Alcohol Dependence 238, 109575.

44. Mehta, N.N., McGillicuddy, F.C., Anderson, P.D., Hinkle, C.C., Shah, R., Pruscino, L., Tabita-Martinez, J., Sellers, K.F., Rickels, M.R., Reilly, M.P., 2009. Experimental Endotoxemia Induces Adipose Inflammation and Insulin Resistance in Humans. Diabetes 59(1), 172–181.

45. Miller, M., Zhan, M., Havas, S., 2005. High Attributable Risk of Elevated C-Reactive Protein Level to Conventional Coronary Heart Disease Risk Factors: The Third National Health and Nutrition Examination Survey. Archives of Internal Medicine 165(18), 2063–2068.

46. Moore, J.X., Chaudhary, N., Akinyemiju, T., 2017. Metabolic Syndrome Prevalence by Race/Ethnicity and Sex in the United States, National Health and Nutrition Examination Survey, 1988-2012. Prev Chronic Dis 14, E24.

47. Nazrul Islam, S.K., Jahangir Hossain, K., Ahmed, A., Ahsan, M., 2002. Nutritional status of drug addicts undergoing detoxification: prevalence of malnutrition and influence of illicit drugs and lifestyle. Br J Nutr 88(5), 507–513.

48. Neale, J., Nettleton, S., Pickering, L., Fischer, J., 2012. Eating patterns among heroin users: a qualitative study with implications for nutritional interventions. Addiction 107(3), 635–641.

49. Nolan, L.J., Scagnelli, L.M., 2007. Preference for sweet foods and higher body mass index in patients being treated in long-term methadone maintenance. Subst Use Misuse 42(10), 1555–1566.

50. Ochalek, T.A., Laurent, J., Badger, G.J., Sigmon, S.C., 2021. Sucrose subjective response and eating behaviors among individuals with opioid use disorder. Drug and Alcohol Dependence 227, 109017.

51. Pappan, N., Awosika, A.O., Rehman, A., 2025. Dyslipidemia, StatPearls. StatPearls Publishing Copyright © 2025, StatPearls Publishing LLC., Treasure Island (FL).

52. Peles, E., Schreiber, S., Sason, A., Adelson, M., 2016. Risk Factors for Weight Gain during Methadone Maintenance Treatment. Substance Abuse 37(4), 613–618.

53. Rosoff, D.B., Charlet, K., Jung, J., Lee, J., Muench, C., Luo, A., Longley, M., Mauro, K.L., Lohoff, F.W., 2019. Association of High-Intensity Binge Drinking With Lipid and Liver Function Enzyme Levels. JAMA Network Open 2(6), e195844–e195844.

54. Ross, R., Neeland, I.J., Yamashita, S., Shai, I., Seidell, J., Magni, P., Santos, R.D., Arsenault, B., Cuevas, A., Hu, F.B., Griffin, B.A., Zambon, A., Barter, P., Fruchart, J.-C., Eckel, R.H., Matsuzawa, Y., Després, J.-P., 2020. Waist circumference as a vital sign in clinical practice: a Consensus Statement from the IAS and ICCR Working Group on Visceral Obesity. Nature Reviews Endocrinology 16(3), 177–189.

55. Suzuki, T., Fukui, S., Shinozaki, T., Asano, T., Yoshida, T., Aoki, J., Mizuno, A., 2025. Lipid Profiles After Changes in Alcohol Consumption Among Adults Undergoing Annual Checkups. JAMA Network Open 8(3), e250583–e250583.

56. Tang, A.M., Forrester, J.E., Spiegelman, D., Flanigan, T., Dobs, A., Skinner, S., Wanke, C., 2010. Heavy injection drug use is associated with lower percent body fat in a multi-ethnic cohort of HIV-positive and HIV-negative drug users from three U.S. cities. Am J Drug Alcohol Abuse 36(1), 78–86.

57. Tishkowski, K., Zubair, M., 2025. Erythrocyte Sedimentation Rate, StatPearls. StatPearls Publishing Copyright © 2025, StatPearls Publishing LLC., Treasure Island (FL).

58. Vallecillo, G., Robles, M.J., Torrens, M., Samos, P., Roquer, A., Martires, P.K., Sanvisens, A., Muga, R., Pedro-Botet, J., 2018. Metabolic Syndrome among Individuals with Heroin use Disorders on Methadone Therapy: Prevalence, Characteristics, and Related Factors. Substance Abuse 39(1), 46–51.

59. Vancampfort, D., Hallgren, M., Mugisha, J., De Hert, M., Probst, M., Monsieur, D., Stubbs, B., 2016. The Prevalence of Metabolic Syndrome in Alcohol Use Disorders: A Systematic Review and Meta-analysis. Alcohol Alcohol 51(5), 515–521.

60. Wakeman, S.E., Larochelle, M.R., Ameli, O., Chaisson, C.E., McPheeters, J.T., Crown, W.H., Azocar, F., Sanghavi, D.M., 2020. Comparative Effectiveness of Different Treatment Pathways for Opioid Use Disorder. JAMA Netw Open 3(2), e1920622.

61. Wei, W., Yang, S., Qiu, Y., Wang, H., Zhao, X., Zhao, Y., Li, Y., Wu, M., Chen, Y., Wang, W., Shi, X., Liu, S., Chen, J., Shen, H., Zhao, D., Su, Y., Shen, C., Yao, Y.S., 2014. CRP gene polymorphism contributes genetic susceptibility to dyslipidemia in Han Chinese population. Mol Biol Rep 41(4), 2335–2343.

62. Welsh, P., Polisecki, E., Robertson, M., Jahn, S., Buckley, B.M., de Craen, A.J., Ford, I., Jukema, J.W., Macfarlane, P.W., Packard, C.J., Stott, D.J., Westendorp, R.G., Shepherd, J., Hingorani, A.D., Smith, G.D., Schaefer, E., Sattar, N., 2010. Unraveling the directional link between adiposity and inflammation: a bidirectional Mendelian randomization approach. J Clin Endocrinol Metab 95(1), 93–99.

63. Wiers, C.E., Martins De Carvalho, L., Hodgkinson, C.A., Schwandt, M., Kim, S.W., Diazgranados, N., Wang, G.J., Goldman, D., Volkow, N.D., 2021. TSPO polymorphism in individuals with alcohol use disorder: Association with cholesterol levels and withdrawal severity. Addict Biol 26(1), e12838.

64. Wiss, D.A., Schellenberger, M., Prelip, M.L., 2019. Rapid Assessment of Nutrition Services in Los Angeles Substance Use Disorder Treatment Centers. Journal of Community Health 44(1), 88–94.

65. Zhu, X., Yu, L., Zhou, H., Ma, Q., Zhou, X., Lei, T., Hu, J., Xu, W., Yi, N., Lei, S., 2018. Atherogenic index of plasma is a novel and better biomarker associated with obesity: a population-based cross-sectional study in China. Lipids in Health and Disease 17(1), 37.

66. Zierle-Ghosh, A., Jan, A., 2025. Physiology, Body Mass Index, StatPearls. StatPearls Publishing Copyright © 2025, StatPearls Publishing LLC., Treasure Island (FL).

