## Supplemental Table 1 for "Adiposity and inflammation mediate altered metabolic profiles in individuals with opioid use disorder"

| **Table 1S. Participant characteristics of individuals with OUD included vs. not included in the medication sub-analysis: number of occurrences (%) or mean (standard deviation)** | | | |
| --- | --- | --- | --- |
|  | **Sub-analysis**  (n= 60) | **Not Included**  (n= 221) | **P- value*** |
| Sex (females) | 23 (38.3%) | 68 (30.8%) | 0.27 |
| Socioeconomic (1-9) | 3.7 (2.2) | 3.9 (2.3) | 0.64 |
| Age (yrs.) | 42.7 (11.8) | 44.9 (12.8) | 0.23 |
| Education (yrs.) | 12.2 (2.4) | 12.4 (2.4) | 0.54 |
| Ethnicity (Hispanic) | 0 (0%) | 5 (2.3%) | 0.12 |
| Race  White  African American  Others/ Unknown | 34 (56.7%)  23 (38.3%)  3 (5%) | 118 (53.4%)  87 (39.4%)  16 (7.2%) | 0.79 |
| Substance Use Disorder  Alcohol  Cannabis  Stimulants | 52 (86.7%)  46 (76.7%)  44 (73.3%) | 187 (84.6%)  159 (71.9%)  157 (71.0%) | 0.89 |
| Smoking (Yes) | 47 (81.0%) | 160 (79.2%) | 0.76 |

| **Table 1S. Participant characteristics by medication for OUD: number of occurrences (%) or mean (standard deviation)** | | | | |
| --- | --- | --- | --- | --- |
|  | No medication (n=11) | Methadone (n=30) | Buprenorphine (n=19) | P- value |
| Race  White  African American  Others/ Unknown | 4 (36%)  7 (64%)  0 (0%) | 19 (63%)  9 (30%)  2 (7%) | 11 (58%)  7 (37%)  1 (5%) | 0.39 |
| Substance use  Alcohol  Cannabis  Stimulants | 10 (91%)  10 (91%)  10 (91%) | 27 (90%)  20 (67%)  21 (70%) | 15 (80%)  16 (84%)  13 (68%) | 0.49  0.17  0.34 |
| Smoking | 5 (45%) | 27 (90%) | 15 (79%) | 0.001 |
| Hispanic | 0 (0%) | 0 (0%) | 0 (0%) | 1.0 |
| Female | 5 (42%) | 14 (48%) | 4 (21%) | 0.17 |
| Socioeconomic (1-9) | 4.6 (2.2) | 3.2 (2.1) | 4.1 (2.3) | 0.19 |
| Age (yrs.) | 36.0 (7.8) | 45.1 (12.2) | 42.7 (12.2) | 0.09 |
| Education (yrs.) | 12.5 (1.5) | 11.9 (2.3) | 12.6 (3.0) | 0.56 |
| Opioid use  Age of onset  Number of years use  Number of years quit | 19.2 (4.9)  12.9 (9.0)  4.5 (3.6) | 21.2 (5.2)  19.5 (11.9)  2.9 (4.7) | 23.1 (9.4)  15.1 (12.3)  2.2 (3.1) | 0.31  0.21  0.31 |
| # with past 12-month dx | 3 (27%) | 25 (83%) | 18 (95%) | <0.001 |

*Independent t-tests were used for numerical and χ^2^ tests were used for categorial variables.

**Alternative hypothesis**

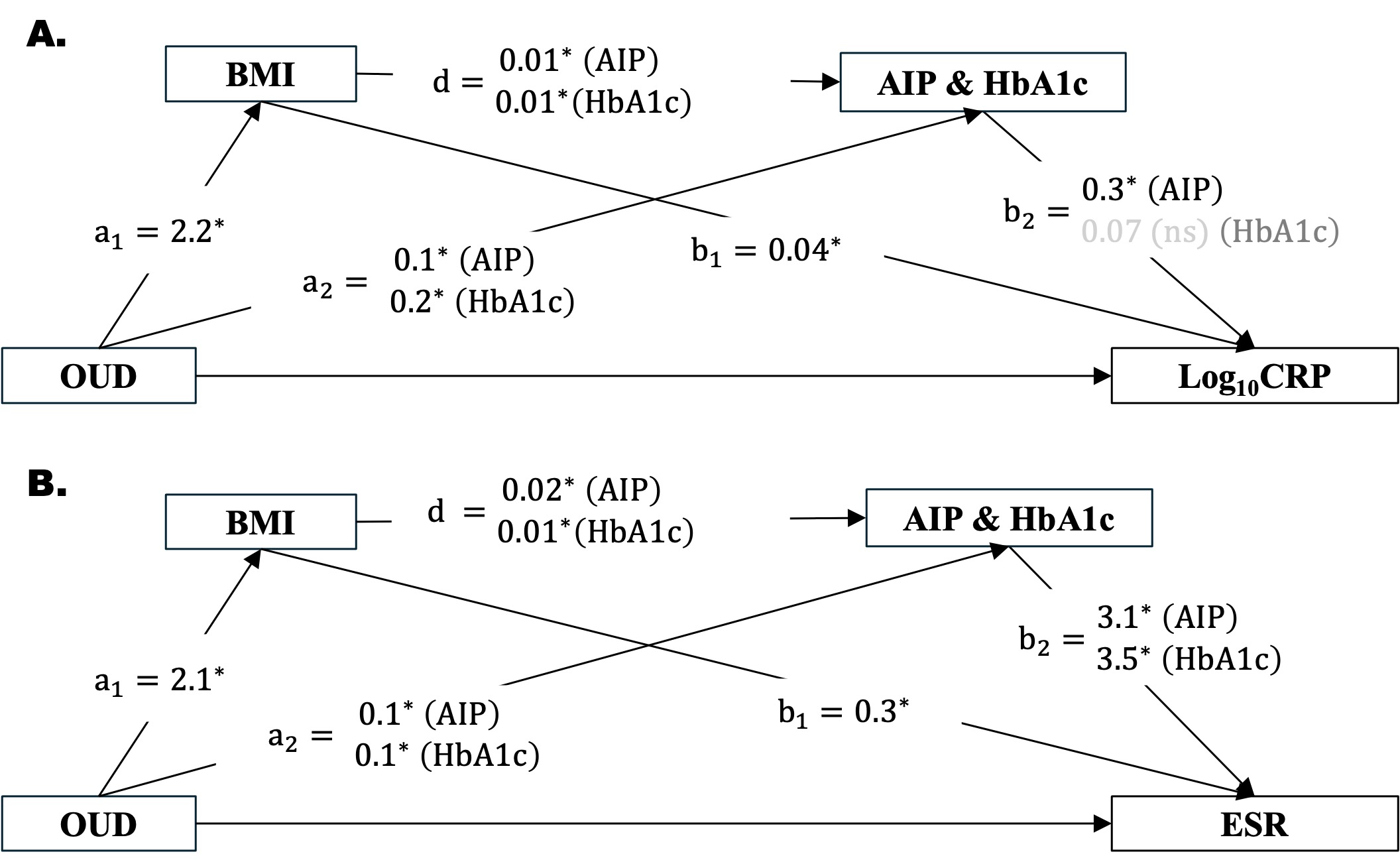

**Figure 1S: Alternative serial mediation model**, in which elevated BMI and metabolic impairment serially mediate OUD-associated elevation in inflammatory markers, (A) log_10_CRP and (B) ESR. Regression coefficients are presented. *, p< 0.05; ns, non-significant. Abbreviations: AIP, atherogenic index of plasma (log10TG/HDL); BMI, body mass index; CRP, C-reactive protein; ESR, erythrocyte sedimentation rate; HbA1c, hemoglobin A1c; OUD, opioid use disorder.

| **Table 2S. Alternative serial mediation model with total and direct effects of OUD on inflammatory markers and indirect effects serially mediated through BMI 🡪 metabolic parameters.** | | | | | |
| --- | --- | --- | --- | --- | --- |
| Metabolic parameter | Inflammatory marker | Total effect | Direct effect | Total indirect effects | Indirect effects  BMI-> metabolic parameters |
| AIP  [99% CI] | Log_10_CRP | 0.16  [0.01, 0.3]* | ns | 0.13  [0.06, 0.2]* | 0.01  [0.002, 0.02]* |
| HbA1c  [99% CI] | Log_10_CRP | 0.16  [0.01, 0.3]* | ns | 0.10  [0.03, 0.2]* | ns |
| AIP  [99% CI] | ESR | 2.7  [-0.03, 5.4]* | ns | 1.2  [0.4, 2.2]* | ns |
| HbA1c  [99% CI] | ESR | 2.8  [0.05, 5.5]* | ns | 1.3  [0.4, 2.5]* | 0.08  [0.003, 0.23]* |
| *Statistical significance, based on 5000 bootstrap sample, 95% CI excluding 0. Abbreviation: AIP, atherogenic index of plasma (log_10_TG/HDL) ; BMI, body mass index, CI, confidence interval, CRP, C-reactive protein; ESR, erythrocyte sedimentation rate; HbA1c, hemoglobin A1c; ns, non-significant; OUD, opioid use disorder. | | | | | |

Examination of alternative mechanism showed significant effects of BMI→ AIP in serially mediating OUD-associated elevation in Log_10_CRP. This aligned with a previous study demonstrating elevated serum levels of proinflammatory cytokines in individuals with dyslipidemia compared to those without [51]. The inflammation response is triggered when triglyceride-rich low-density lipoprotein cholesterol accumulated in the arterial endothelium, undergoes oxidative modification, and subsequently act as substrates for immune cells recognition and activation [52]. In contrast, HDL-cholesterol exerts anti-inflammatory effects by facilitating cholesterol efflux from the arterial endothelial cells, inhibiting LDL oxidation, and reducing the recruitment of inflammatory cells [53]. Furthermore, our finding of a significant BMI→ HbA1c effect in mediating elevated ESR in individuals with OUD is consistent with extensive evidence showing the pro-inflammatory effects of glucose [54, 55].

**Metabolic and inflammatory alterations in OUD, AUD, and their co-occurrence**

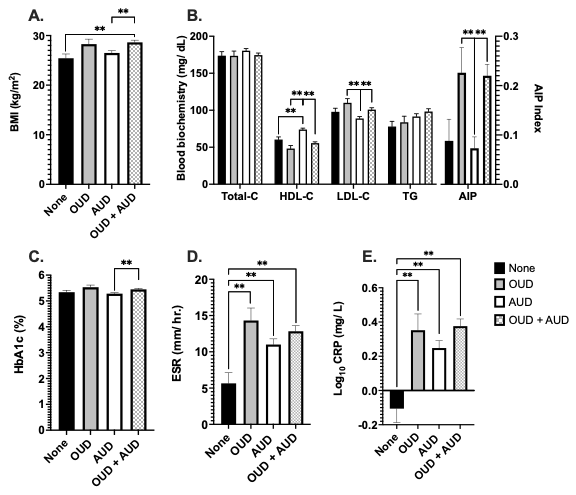

**Figure 2S** **Metabolic and inflammatory parameters** **in controls and individuals with OUD only, AUD only, and OUD+AUD.** (A) BMI, (B) lipid profiles, (C) HbA1c, (D) ESR, and (E) log-transformed CRP, adjusting for stimulant use disorder, cannabis use disorder, and smoking status, in individuals with none (n= 75), OUD only (n=41), AUD only (n= 171), and OUD+AUD (n=239). Means ± SE. **Indicates significance p< 0.05. Abbreviations: AIP, atherogenic index of plasma (log10TG/HDL); AUD, alcohol use disorder; BMI, body mass index, CRP, C-reactive protein; ESR, erythrocyte sedimentation rate; HbA1c, hemoglobin A1c; HDL-C, high-density lipoprotein cholesterol; LDL-C, low-density lipoprotein cholesterol; TG, triglycerides; Total-C, total cholesterol; OUD, opioid use disorder.

Multivariate comparison between individuals with OUD only, AUD only, OUD+AUD, and controls, adjusting for stimulant and cannabis use disorder and smoking status, revealed a significant effect on the combined set of dependent variables (MANCOVA F_27, 1376_= 5.6, p< 0.001, Wilks’ λ= 0.7, η_p_^2^=0.1) (**Figure 2S**). Follow-up univariate tests showed significant group effects on BMI (**Figure 2SA**, F_3, 479_= 4.7, p= 0.003 η_p_^2^=0.03), such that it was higher in individuals with OUD+AUD compared to controls and individuals with AUD only. There were significant effects of group in altering lipid profile (**Figure 2SB**), including HDL-C (F_3, 479_= 21.2, p< 0.001, η_p_^2^=0.1), LDL-C (F_3, 479_= 5.9, p< 0.001, η_p_^2^=0.04), and AIP (F_3, 479_= 7.0, p< 0.001, η_p_^2^=0.05). *Post-hoc* analysis showed higher LDL-C and AIP in individuals with OUD only and OUD+AUD compared to those with AUD only, and higher HDL-C in individuals with AUD compared to all other groups (p’s< 0.05). There were no significant differences in total cholesterol (F_3, 479_= 1.0, p= 0.4, η_p_^2^=0.006) or triglyceride levels (F_3, 479_= 2.2, p=0.08, η_p_^2^=0.01). There was a significant group difference on HbA1c (**Figure 2SC**, F_3, 479_= 3.9, p= 0.000, η_p_^2^=0.02) with higher levels in individuals with OUD+AUD than those with AUD only. Moreover, there were significant group differences in inflammatory markers, ESR (**Figure 2SD**, F_3, 479_= 6.2, p< 0.001, η_p_^2^=0.04) and log_10_CRP (**Figure 2SE**, F_3, 479_= 7.9, p< 0.001, η_p_^2^=0.05) and *post-hoc* analysis showed lower levels in controls than all other groups.

**Metabolic and inflammatory parameters in individuals with OUD with vs. without a current diagnosis within the past 12 months**

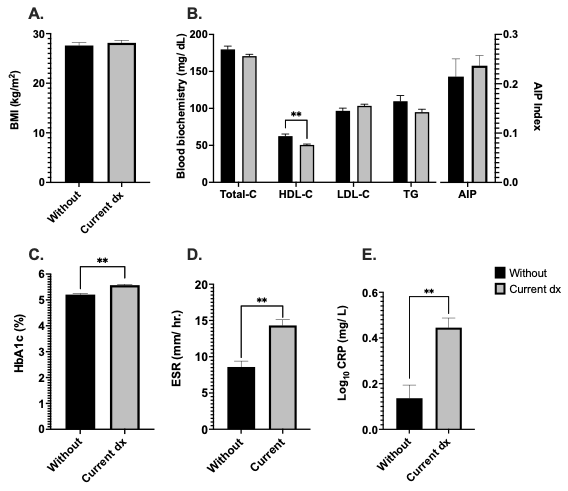

**Figure 3S** **Metabolic and inflammatory parameters** **in individuals with OUD without vs. with current diagnosis within the past 12 months.** (A) BMI, (B) lipid profiles, (C) HbA1c, (D) ESR, and (E) log-transformed CRP. Means ± SE. **Indicates significance p< 0.05. Abbreviations: AIP, atherogenic index of plasma (log10TG/HDL); BMI, body mass index, CRP, C-reactive protein; ESR, erythrocyte sedimentation rate; HbA1c, hemoglobin A1c; HDL-C, high-density lipoprotein cholesterol; LDL-C, low-density lipoprotein cholesterol; TG, triglycerides; Total-C, total cholesterol

Compared to individuals who did not use opioids in the past 12 months, those who have did had lower HDL-C [t_121.2_= 3.8, p< 0.001], HbA1c [t_277_= -6.2, p< 0.001], log_10_CRP [t_278_= -4.2, p< 0.001], and ESR [t_226.2_= -5.1, p< 0.001]. There were no significant differences on BMI [t_214.6_= -0.7, p= 0.5], total-C [t_279_= 2.0, p= 0.07], LDL-C [t_276_= -1.6, p= 0.1], triglycerides [t_279_= 1.9, p= 0.06], and AIP [t_279_= -0.6, p= 0.6] (Figure 3S).

**Table 2S. Pearson correlations between metabolic and inflammatory parameters.**

|  | BMI | Total- C | HDL- C | LDL- C | TG | Hgb A1C | ESR | Log_10_ CRP |
| --- | --- | --- | --- | --- | --- | --- | --- | --- |
| BMI | - | .072 | -.296^**^ | .247^**^ | .177^**^ | .143^**^ | .220^**^ | .442^**^ |
| Total-C | .072 | - | .356^**^ | .772^**^ | .306^**^ | .017 | .095^*^ | .011 |
| HDL-C | -.296^**^ | .356^**^ | - | -.269^**^ | -.261^**^ | -.144^**^ | -.195^**^ | -.263^**^ |
| LDL-C | .247^**^ | .772^**^ | -.269^**^ | - | .299^**^ | .106^*^ | .233^**^ | .148^**^ |
| TG | .177^**^ | .306^**^ | -.261^**^ | .299^**^ | - | .066 | .071 | .189^**^ |
| Hgb A1C | .143^**^ | .017 | -.144^**^ | .106^*^ | .066 | - | .209^**^ | .147^**^ |
| ESR | .220^**^ | .095^*^ | -.195^**^ | .233^**^ | .071 | .209^**^ | - | .462^**^ |
| Log_10_ CRP | .442^**^ | .011 | -.263^**^ | .148^**^ | .189^**^ | .147^**^ | .462^**^ | - |
| *Correlation is significant at p< 0.05; **Correlation is significant at p< 0.001. | | | | | | | | |
